# Geographic variation in loss to follow-up from HIV care in Tanzania and its association with pharmacy refill adherence in routine programme data

**DOI:** 10.64898/2026.03.04.26347648

**Authors:** Meshack D. Lugoba, Raphael Z. Sangeda, Lucas De Vrieze, Hillary Mushi, Ritah F. Mutagonda, James Mwakyomo, Veryeh Sambu, Prosper Njau

## Abstract

**Background:** Sustained retention in HIV care is essential for achieving durable viral suppression and controlling the HIV epidemic. Loss to follow-up (LTFU) remains a persistent challenge in sub-Saharan Africa and shows substantial geographic variation. However, nationally representative analyses of routine monitoring data remain limited. Pharmacy refill data provide a scalable and objective approach for identifying individuals at risk of disengaging from care. We assessed the magnitude, spatial distribution, and predictors of LTFU among people living with HIV (PLHIV) receiving antiretroviral therapy (ART) across 26 mainland regions of Tanzania.

**Methods:** We conducted a retrospective cohort analysis using routinely collected program data from the National Care and Treatment Clinic (CTC-2) database of PLHIV receiving ART in Tanzania between 2017 and 2021. LTFU was defined as no recorded clinic visit for ≥180 days after the last scheduled appointment, consistent with monitoring definitions used by the National AIDS and Sexually Transmitted Infections Control Programme (NASHCoP). Pharmacy refill adherence was calculated longitudinally and categorized as good (≥85%) or poor (<85%). Regional and district-level patterns were visualized using geospatial mapping. Multivariable logistic regression models were used to identify predictors of LTFU.

**Results:** A total of 52,828 PLHIV were included in the study, representing all 26 mainland regions of Tanzania. Overall, 26.6% were classified as LTFU during follow-up, with marked regional variation. The highest proportional LTFU was observed in Dar es Salaam (33.2%), followed by Njombe (32.9%) and Geita (32.7%), while the lowest was recorded in Mwanza (19.1%) and Iringa (20.3%). Good pharmacy refill adherence (≥85%) was strongly associated with lower odds of LTFU and remained the most robust independent predictor after adjustment (adjusted odds ratio [aOR] 0.34; 95% confidence interval [CI] 0.32-0.35). District-level analyses revealed substantial within-region heterogeneity, identifying localized clusters of elevated attrition not apparent in regional aggregates.

**Conclusion:** LTFU remains a major challenge to sustaining effective ART delivery in Tanzania. Pharmacy refill adherence may serve as a practical early indicator for identifying individuals at risk of disengagement from HIV care. Integrating refill-based monitoring with spatially informed analysis may support targeted retention strategies within routine HIV treatment programs.

## Introduction

The scale-up of antiretroviral therapy (ART) has transformed HIV infection into a manageable chronic condition, substantially reducing the morbidity and mortality worldwide. Eastern and Southern Africa continue to bear the highest global HIV burden despite major progress in treatment coverage and viral suppression [1–3]. Tanzania has rapidly expanded ART coverage over the past decade, alongside a national transition to dolutegravir-based regimens and the adoption of differentiated service delivery models (SDS)[4].

However, achieving sustained viral suppression depends not only on ART initiation but also on long-term retention of care. Loss to follow-up (LTFU) undermines individual clinical outcomes and weakens population-level HIV control by increasing the risk of viral rebound, drug resistance, and onward transmission [5–8]. In sub-Saharan Africa, 20-40% of patients disengage from care within the first few years of ART initiation, with wide variation across countries and regions [9–11].

In Tanzania, previous studies have documented substantial LTFU in both urban and rural ART programmers, with reported rates ranging from approximately 20% to over 35%, depending on the region, follow-up duration, and patient characteristics [12–15]. Recent subnational analyses have further demonstrated that disengagement dynamics differ by epidemiological and geographic contexts. In Manyara, a geographically dispersed region characterized by semi-pastoral mobility patterns, pharmacy refill adherence was shown to predict subsequent loss to follow-up, underscoring the role of structural access constraints and movement dynamics in shaping retention outcomes [16]. In contrast, in Njombe, a high-prevalence region with long-standing ART cohorts, predictors of LTFU were more strongly associated with demographic and clinical characteristics within a mature treatment programmed [17]. While these findings highlight regional heterogeneity, whether these patterns reflect localized phenomena or broader national trends has not been systematically examined.

Monitoring adherence and identifying individuals at risk of disengaging from care are critical programmatic priorities. Self-reported adherence and appointment-based measures are widely used but are subject to recall and social desirability biases. Pharmacy refill adherence has emerged as an objective, low-cost, and scalable alternative, with strong evidence from Tanzania and other settings demonstrating its ability to predict viral failure and treatment interruption [18–21]. However, the performance of refill adherence as a predictor of LTFU within national routine programme data has not been comprehensively evaluated across diverse regional contexts.

Therefore, this study aimed to (i) quantify the magnitude of LTFU among people living with HIV (PLHIV) receiving ART across all 26 mainland regions of Tanzania, (ii) characterize regional and district-level heterogeneity in LTFU using spatial analysis, and (iii) evaluate the predictive value of pharmacy refill adherence for LTFU within the routinely collected national programme data. By integrating evidence from both high- and low-prevalence settings, this analysis provides nationally relevant insights to inform geographically targeted and differentiated retention strategies.

## Methods

### Study population and cohort construction

The national Care and Treatment Clinic (CTC-2) database contains routine program records for individuals receiving ART in Tanzania. During the study period (January 2017 to December 2021), the database included more than two million patient records. To obtain a computationally manageable but nationally representative analytic dataset, a simple random sample of approximately 60,000 individuals with at least one recorded ART refill or clinic visit during the study period was drawn from the national database.

Eligibility criteria were then applied to ensure valid longitudinal follow-up. Records were excluded if they lacked key identifiers required for linkage across visits, including missing or inconsistent patient identifiers or first recorded clinic visit dates. Additional exclusions were applied to records without sufficient follow-up information to determine retention status. After these exclusions, 52,828 individuals living with HIV with valid longitudinal data were retained for analysis, representing patients from all 26 mainland regions of Tanzania. The geographic distribution of included individuals is provided in Supplementary Table S1.

The analytic cohort retained representation from all mainland regions and districts captured in the national registry during the study period. Because the sample was drawn using simple random sampling from the national database, the geographic and demographic structure of the cohort reflects that of the underlying ART population recorded in the CTC-2 registry.

### Sampling strategy

To ensure the analytic dataset remained nationally representative, a simple random sampling procedure was used to select individuals from the national registry, without stratification by facility, region, or demographic characteristics. Because each individual in the registry had an equal probability of selection, the resulting analytic cohort preserves the national geographic distribution and demographic structure of the underlying population while maintaining computational efficiency for repeated longitudinal analyses.

### Definition of loss to follow-up

Loss to follow-up (LTFU) was defined as the absence of any recorded clinic visit during the 180 days preceding the end of the observation window (31 December 2021), based on documented visit dates in the registry. This operational definition relies on recorded service encounters rather than registry outcome fields and provides a standardized measure of disengagement from care within the study period.

### Use of registry status fields

The CTC-2 registry also includes a program-reported final status field (e.g., attending clinic, missing appointments, lost to follow-up, transfer out, death) that is updated through facility-level tracing activities. Because these updates may occur asynchronously and inconsistently across facilities, registry status fields were not used to define the primary LTFU outcome in this analysis. Instead, visit-date gaps were used to derive a standardized measure of disengagement from care. The distribution of registry-reported final status is presented descriptively in the supplementary material.

### Pharmacy refill adherence

Pharmacy refill adherence was calculated longitudinally by measuring medication possession across consecutive refill intervals and expressed as the percentage of days covered during follow-up. Adherence was categorized as good (≥85%) or poor (<85%), consistent with prior validation studies conducted in Tanzania and similar, resource-limited settings.

### Covariates

Covariates included age at first recorded clinic visit, gender, marital status, region and district of residence, year of ART initiation and duration of follow-up. Viral load measurements and viral suppression status were included when applicable. Geographic variables were harmonized using standardized region and district coding to ensure consistency across facilities and over time.

### Statistical analysis

Descriptive statistics were used to summarize baseline characteristics, adherence, and LTFU. Regional- and district-level patterns of LTFU and adherence were visualized using geospatial mapping in R. Aggregated regional and district estimates were calculated from individual-level data and linked to administrative boundary shapefiles to generate thematic maps. Logistic regression was used to evaluate factors associated with LTFU. Univariate models were first fitted for each predictor, followed by a multivariable logistic regression model including demographic and programmatic variables. The multivariable logistic regression model included only categorical covariates (pharmacy refill adherence category, gender, marital status, age group, and region). Age was modeled using predefined age categories rather than continuous age to avoid collinearity and facilitate interpretation in relation to programmatically relevant population groups. The results are presented as odds ratios (ORs) or adjusted odds ratios (aORs) with 95% confidence intervals (CIs). Statistical significance was assessed using a two-sided p-value of <0.05. than 0.05.

### Ethical considerations

Ethical approval for this study was obtained from the Muhimbili University of Health and Allied Sciences Institutional Review Board (MUHAS-REC-02-2022-972; issued 09 February 2022). The analysis used secondary, de-identified routine program data extracted from the national HIV Care and Treatment Clinic (CTC-2) database. As the study involved no direct participant contact and posed minimal risk, the Institutional Review Board waived the requirement for individual informed consent. All data were handled in accordance with national data protection and confidentiality guidelines.

## Results

### Cohort characteristics

A total of 52,828 people living with HIV (PLHIV) receiving antiretroviral therapy between 2017 and 2021 were included in the analysis (Table 1). Females comprised 34,592 (65.5%) of the cohort, whereas males comprised 18,236 (34.5%). Most participants were adults aged 29-69 years (39,518; 74.8%). Young adults aged 19-28 years accounted for 9,079 (17.2%), adolescents aged 11-18 years for 1,680 (3.2%), children aged 0-10 years for 1,922 (3.6%), and elderly individuals aged ≥70 years for 623 (1.2%). Age group information was missing for six individuals (<0.1%). Nearly half of the participants were married (26,023; 49.3%), followed by single (11,057; 20.9%), divorced or separated (3,774; 7.1%), widowed (2,737; 5.2%), and cohabiting (642; 1.2%). Marital status was unknown or not recorded for 8,595 individuals (16.3%). Participants were drawn from all 26 mainland regions of Tanzania. The largest contributions were from Dar es Salaam (6,866; 13.0%), Mbeya (4,010; 7.6%), and Mwanza (4,000; 7.6%), followed by Kagera (3,110; 5.9%), Tabora (2,737; 5.2%), Geita (2,625; 5.0%), and Shinyanga (2,631; 5.0%). Smaller regional contributions were observed in Manyara (594; 1.1%), Katavi (920; 1.7%), Kigoma (816; 1.5%), and Lindi (811; 1.5%). The geographic distribution of participants across regions is presented in Supplementary Table S1.

**Table 1:**
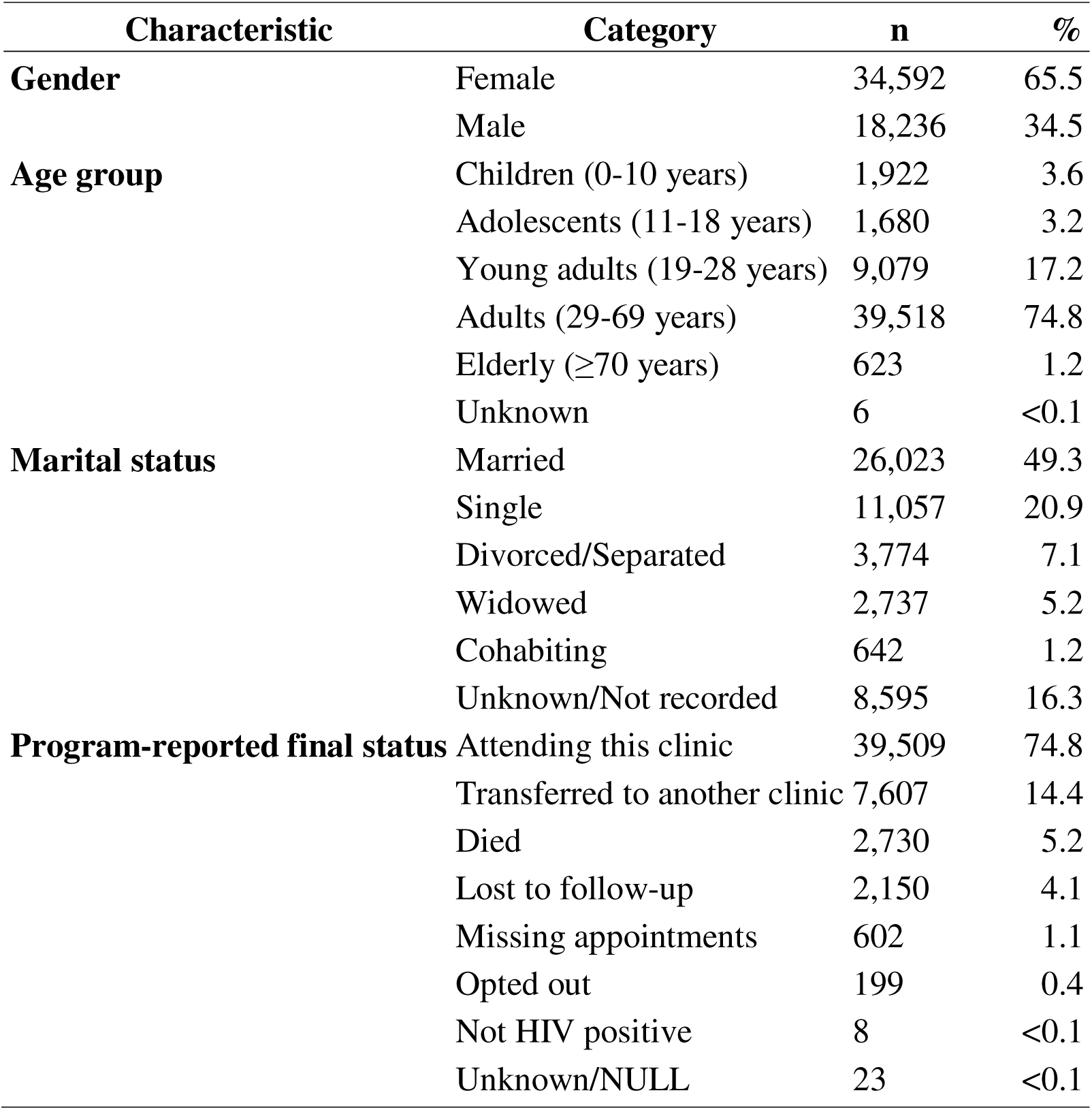
Baseline characteristics of people living with HIV receiving antiretroviral therapy in Tanzania, 2017-2021 (N = 52,828)

| **Characteristic** | **Category** | **n** | **%** |
| --- | --- | --- | --- |
| **Gender** | Female | 34,592 | 65.5 |
|  | Male | 18,236 | 34.5 |
| **Age group** | Children (0-10 years) | 1,922 | 3.6 |
|  | Adolescents (11-18 years) | 1,680 | 3.2 |
|  | Young adults (19-28 years) | 9,079 | 17.2 |
|  | Adults (29-69 years) | 39,518 | 74.8 |
| | Elderly ($\geq 70$ years) | 623 | 1.2 |
|  | Unknown | 6 | <0.1 |
| **Marital status** | Married | 26,023 | 49.3 |
|  | Single | 11,057 | 20.9 |
|  | Divorced/Separated | 3,774 | 7.1 |
|  | Widowed | 2,737 | 5.2 |
|  | Cohabiting | 642 | 1.2 |
|  | Unknown/Not recorded | 8,595 | 16.3 |
| **Program-reported final status** | Attending this clinic | 39,509 | 74.8 |
|  | Transferred to another clinic | 7,607 | 14.4 |
|  | Died | 2,730 | 5.2 |
|  | Lost to follow-up | 2,150 | 4.1 |
|  | Missing appointments | 602 | 1.1 |
|  | Opted out | 199 | 0.4 |
|  | Not HIV positive | 8 | <0.1 |
|  | Unknown/NULL | 23 | <0.1 |

### Pharmacy refill adherence and loss to follow-up

Overall, 34,257 of 52,828 participants (64.8%) demonstrated good pharmacy refill adherence (≥85%), 18,158 (34.4%) had poor adherence (<85%), and 413 (0.8%) had missing adherence data (Table 2).

**Table 2:**
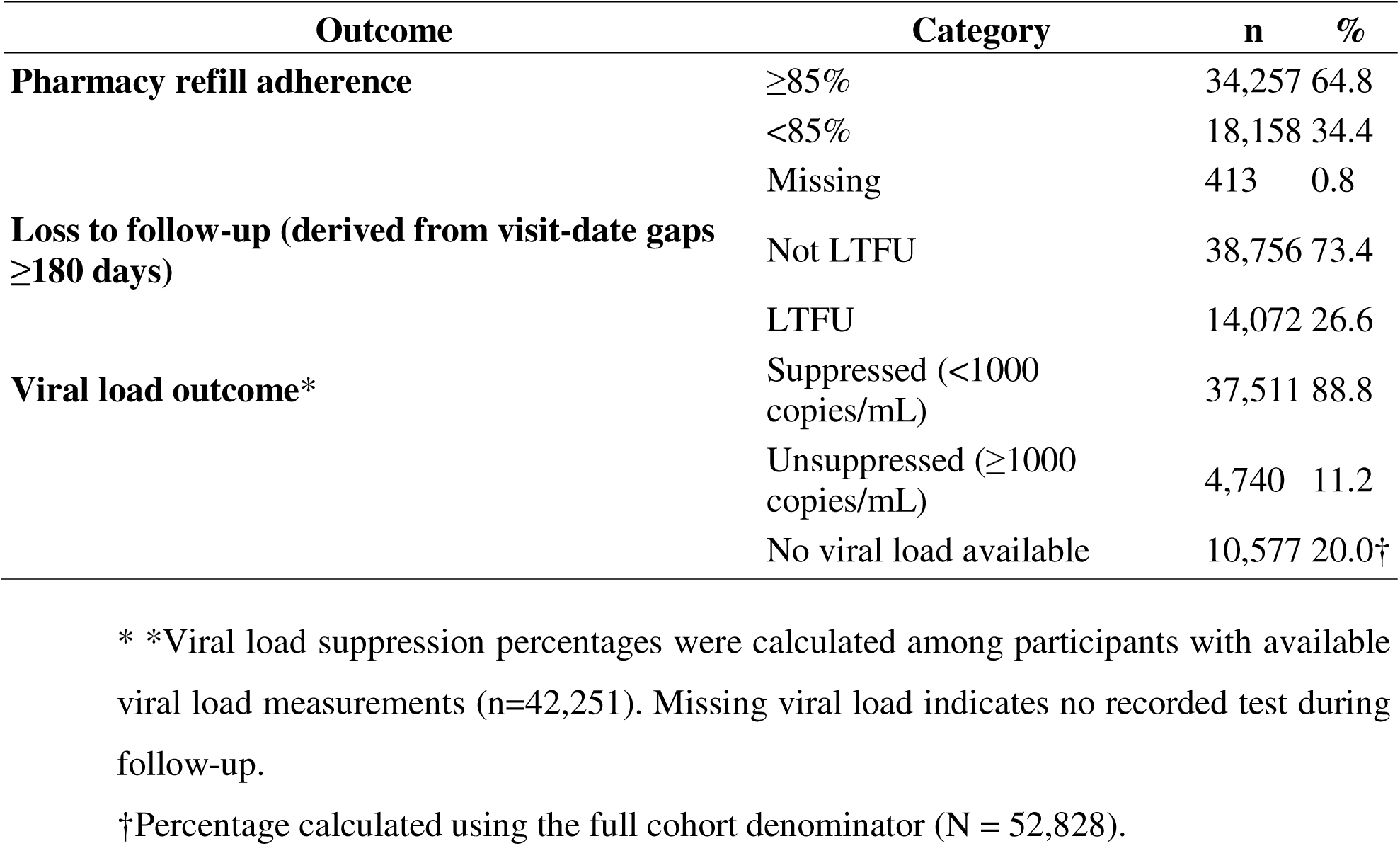
National pharmacy refill adherence, viral suppression, and loss to follow-up outcomes among PLHIV receiving ART in Tanzania, 2017-2021 (N = 52,828).

Loss to follow-up (LTFU), defined as the absence of any recorded clinic visit during the 180 days preceding the end of the observation window, occurred in 14,072 participants (26.6%), whereas 38,756 (73.4%) remained in care according to this operational definition. Viral load measurements were available for 42,251 participants (80.0%), of whom 37,511 (88.8%) achieved viral suppression (<1000 copies/mL). Outcome data availability differed across analyses: LTFU status was available for the entire cohort (n = 52,828), pharmacy refill adherence for 52,415 participants, and viral load measurements for 42,251 participants.

### Regional variation in adherence and loss to follow-up

Substantial regional heterogeneity in loss to follow-up (LTFU) and pharmacy refill adherence was observed across mainland Tanzania (Table 3; Figure 1). The highest regional LTFU was recorded in Dar es Salaam (33.2%), followed by Njombe (32.9%), Kigoma (32.8%), Geita (32.7%), and Katavi (32.4%). In contrast, the lowest LTFU rates were observed in Mwanza (19.1%), Iringa (20.3%), and Mara (20.5%).

**Table 3:**
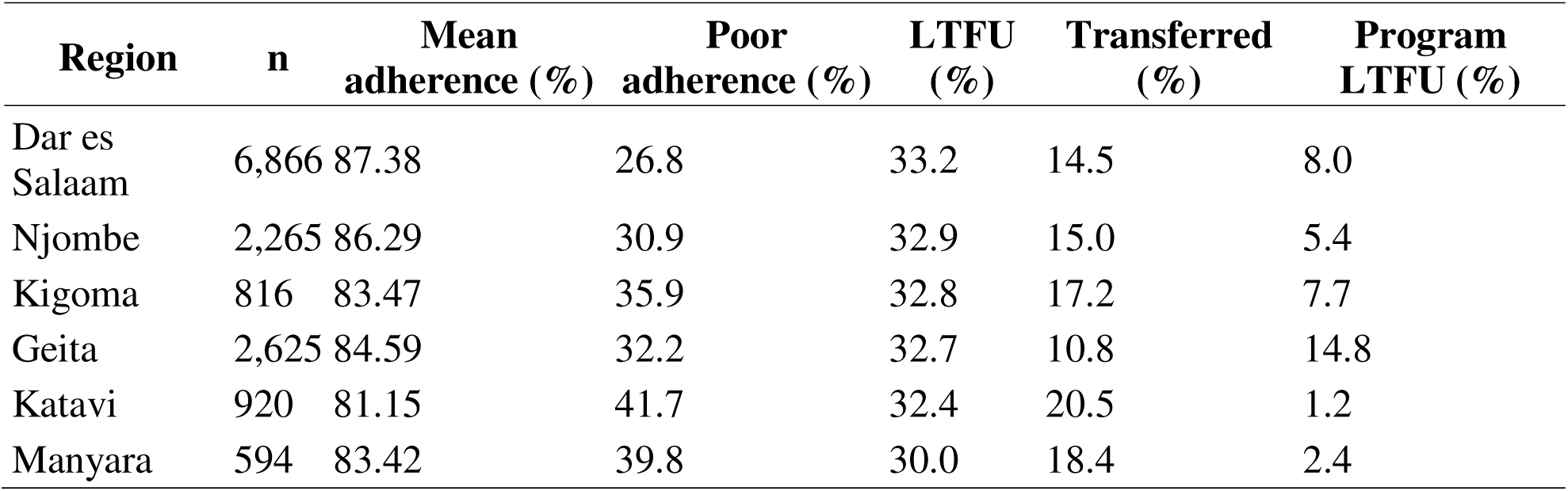
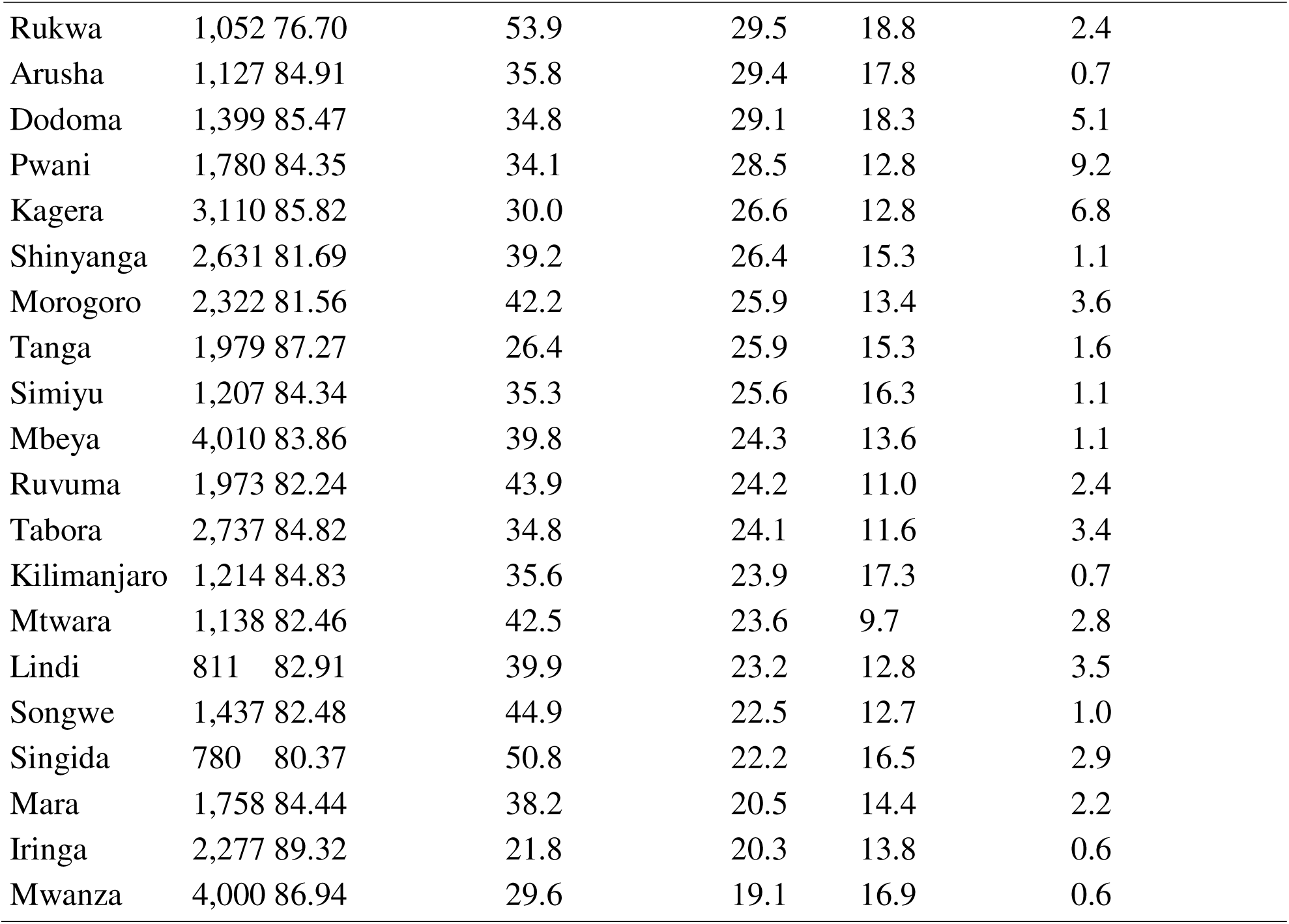
Regional distribution of pharmacy refill adherence and derived loss to follow-up among PLHIV receiving ART in mainland Tanzania, 2017-2021 (N = 52,828) Mean refill adherence, proportion with poor adherence (<85%), and proportion classified as LTFU (≥180 days without a recorded clinic visit) were aggregated from individual-level data across the 26 mainland regions. Program indicators for transfers and registry-reported LTFU are shown for descriptive comparison.

**Figure 1:**
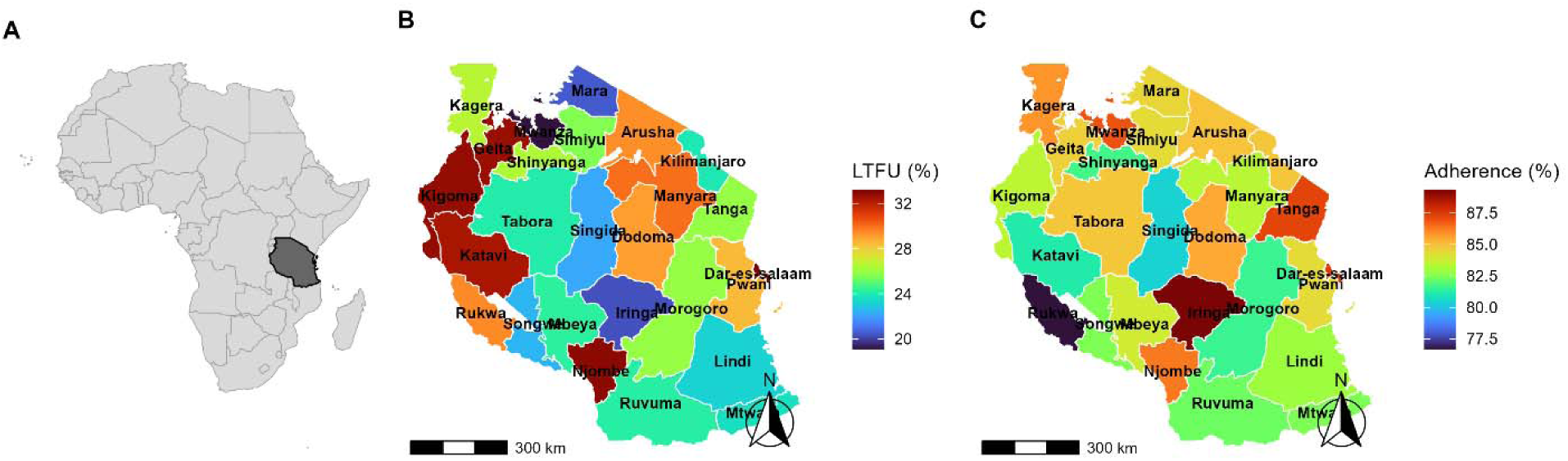
Regional distribution of loss to follow-up (LTFU), and pharmacy refill adherence among people living with HIV receiving antiretroviral therapy in Tanzania, 2017-2021. Panel A shows the location of mainland Tanzania in Africa. Panel B presents the regional distribution of LTFU (≥180 days without recorded clinic visits). Panel C shows the mean pharmacy refill adherence by region, expressed as a percentage of days covered during follow-up. The analyses were restricted to mainland Tanzania, excluding Zanzibar. Darker shading represents higher LTFU (Panel B) or higher adherence (Panel C).

Mean pharmacy refill adherence ranged from 89.3% in Iringa to 76.7% in Rukwa. Regions with higher proportions of poor adherence generally had higher LTFU rates. For example, Rukwa (53.9% poor adherence) and Singida (50.8% poor adherence) also demonstrated relatively high LTFU rates compared with regions with stronger adherence profiles.

The proportion of patients recorded as transferred to another clinic and the program-reported LTFU status recorded in the registry are shown for comparison (Table 3).

### District-level patterns in extreme regions

District-level mapping revealed substantial within-region variation in LTFU (Figure 2; Supplementary Table S2). In Dar es Salaam, district-level LTFU ranged from 26.9% in Temeke to 37.4% in Kigamboni, with similarly elevated values observed in Ilala (36.6%) and Ubungo (36.8%), while Kinondoni showed somewhat lower levels (32.9%).

**Figure 2:**
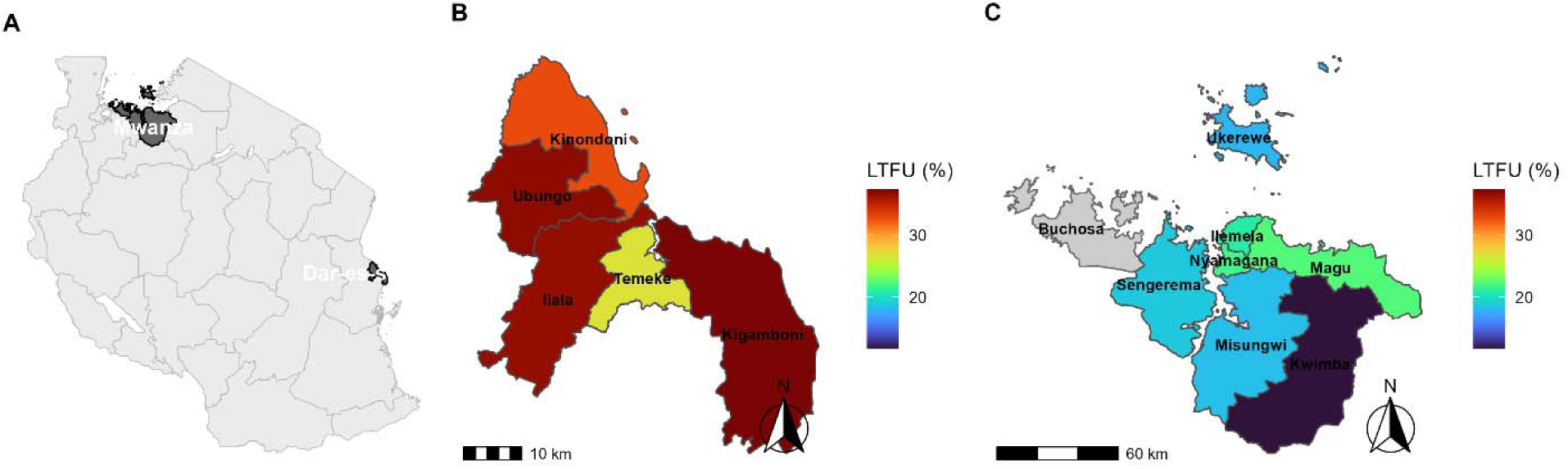
District-level loss to follow-up (LTFU), in regions with the highest and lowest regional LTFU in Tanzania. Panel A highlights the two mainland regions with the highest (Dar es Salaam) and lowest (Mwanza) regional LTFU identified from individual-level analyses. Panels B and C show district-level LTFU in Dar es Salaam and Mwanza, respectively. District-level LTFU was calculated as the proportion of individuals classified as lost to follow-up (≥180 days without recorded clinical visits). Districts with insufficient data are shaded in grey. A shared colour scale is used across Panels B and C to facilitate direct comparison.

In contrast, Mwanza exhibited lower, more homogeneous district-level LTFU rates, ranging from 11.7% in Kwimba to 22.3% in Magu, with most districts clustering around 18-22%.

District-level adherence and LTFU estimates for these regions are presented in Supplementary Table S2.

### LTFU by ART initiation cohort

LTFU varied by year of the first recorded clinic visit. Higher proportions of LTFU were observed among individuals whose first recorded visit occurred in earlier calendar years, with progressively lower proportions among those entering care more recently (Supplementary Figure S1; Supplementary Table S3).

### Predictors of LTFU

In univariate logistic regression analyses (Table 4), pharmacy refill adherence was strongly associated with LTFU. Participants with good refill adherence (≥85%) had substantially lower odds of LTFU compared with those with poor adherence (<85%) (OR 0.35; 95% CI 0.34-0.36; p < 0.001). Increasing age was also associated with slightly lower odds of LTFU (OR 0.99 per year; 95% CI 0.987-0.990; p < 0.001).

**Table 4:**
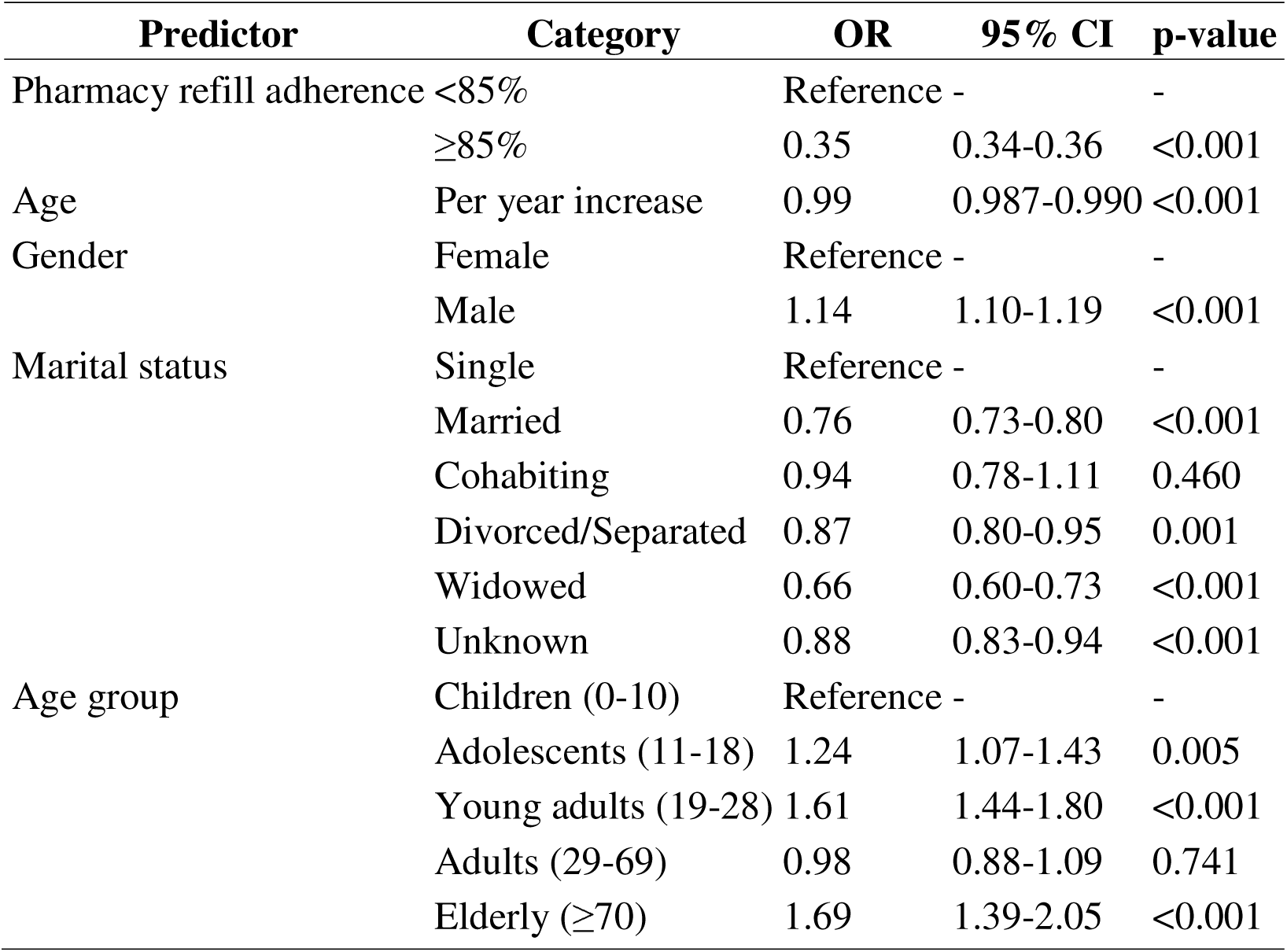
Univariate logistic regression analysis of factors associated with loss to follow-up (LTFU) among PLHIV receiving ART in Tanzania. Odds ratios (ORs) and 95% confidence intervals are presented for demographic, clinical, and regional variables associated with LTFU.

Male gender was associated with higher odds of LTFU compared with female participants (OR 1.14; 95% CI 1.10-1.19; p < 0.001). Marital status also showed associations with LTFU, with married individuals demonstrating lower odds compared with other marital categories. Age-group analysis showed increased odds of LTFU among adolescents (11-18 years), young adults (19-28 years), and elderly individuals (≥70 years) compared with children aged 0-10 years.

Substantial geographic variation in LTFU was also observed. Several regions showed higher odds of LTFU compared with the reference region, including Dar es Salaam, Njombe, Kigoma, Geita, Katavi, and Manyara.

In the multivariable logistic regression model (Table 5), pharmacy refill adherence remained the strongest independent predictor of LTFU. Participants with good adherence (≥85%) had significantly lower odds of LTFU compared with those with poor adherence (aOR 0.34; 95% CI 0.32-0.35; p < 0.001).

**Table 5:** Multivariable logistic regression model of predictors of loss to follow-up among PLHIV receiving ART in Tanzania. Adjusted odds ratios (aORs) with 95% confidence intervals are presented for all variables retained in the final multivariable model.

| Predictor | Category | aOR | 95% CI | p-value |
| --- | --- | --- | --- | --- |
| Pharmacy refill adherence | <85% | Reference | - | - |
|  | ≥85% | 0.34 | 0.32-0.35 | <0.001 |
| Gender | Female | Reference | - | - |
|  | Male | 1.22 | 1.17-1.27 | <0.001 |
| Marital status | Single | Reference | - | - |
|  | Married | 0.81 | 0.76-0.85 | <0.001 |
|  | Cohabiting | 0.97 | 0.81-1.17 | 0.756 |
|  | Divorced/Separated | 0.99 | 0.91-1.08 | 0.872 |
|  | Widowed | 0.81 | 0.73-0.90 | <0.001 |
|  | Unknown | 0.89 | 0.83-0.95 | <0.001 |
| Age group | Children (0-10) | Reference | - | - |
|  | Adolescents (11-18) | 1.28 | 1.09-1.49 | 0.002 |
|  | Young adults (19-28) | 1.76 | 1.56-1.98 | <0.001 |
|  | Adults (29-69) | 1.15 | 1.03-1.29 | 0.015 |
|  | Elderly (≥70) | 2.11 | 1.72-2.59 | <0.001 |

Male gender remained independently associated with higher odds of LTFU (aOR 1.22; 95% CI 1.17-1.27; p < 0.001). Age-group differences persisted after adjustment, with adolescents (aOR 1.28), young adults (aOR 1.76), and elderly individuals (aOR 2.11) showing higher odds of LTFU compared with children aged 0-10 years.

Regional variation remained evident after adjustment for demographic and adherence factors. Higher adjusted odds of LTFU were observed in several regions, including Dar es Salaam (aOR 2.27), Geita (aOR 2.12), Njombe (aOR 2.21), Kigoma (aOR 2.04), and Manyara (aOR 1.74). In contrast, several other regions showed more modest increases.

The adjusted odds ratios for the different regions are shown in Supplementary Table S4.

## Discussion

Using a large, nationally representative sample drawn from routine HIV programme data, this study quantified the magnitude and geographic distribution of loss to follow-up (LTFU) among people receiving antiretroviral therapy (ART) in mainland Tanzania and identified pharmacy refill adherence as a strong predictor of disengagement from care. Substantial heterogeneity in retention was observed across regions and districts, indicating that programme performance varies geographically rather than uniformly across the country. By analyzing patient-level routine monitoring data at a national scale, these findings extend evidence beyond single-facility or regional cohorts and demonstrate the potential of routinely collected programme data to identify populations at elevated risk of attrition.

A central finding of this study was the strong and consistent association between pharmacy refill adherence and LTFU. Individuals with good refill adherence (≥85%) had substantially lower odds of disengagement from care compared with those with poorer adherence. Because refill adherence reflects medication pickup behavior routinely captured in clinical systems, it may detect early disengagement before prolonged absence from clinic visits becomes evident. Similar associations between refill gaps and adverse outcomes, including disengagement and virological failure, have been documented across multiple low- and middle-income settings [7–12]. Tanzanian studies have also shown that refill adherence can identify individuals at increased risk of adverse outcomes more reliably than self-reported measures [9,10]. The present analysis demonstrates that this predictive relationship persists when applied within a nationally representative routine programme dataset.

Marked geographic heterogeneity in LTFU indicates that factors beyond underlying HIV prevalence influence disengagement from care. Regions such as Dar es Salaam, Njombe, and Kigoma showed some of the highest proportions of individuals classified as LTFU. In contrast, several geographically dispersed regions showed high proportional attrition, suggesting structural and access-related barriers to continuity of care. These patterns are consistent with prior region-specific studies in Tanzania. In Manyara, a geographically dispersed setting characterized by population mobility, refill-based adherence trajectories predicted subsequent disengagement, highlighting continuity-of-care challenges [16]. In Njombe, where a mature treatment programme with long-standing ART cohorts was in place, demographic and clinical predictors played a greater role in retention outcomes [17]. The present district-level mapping further identified sub-regional pockets of elevated attrition, demonstrating the value of granular spatial analyses for geographically targeted programme interventions [13,21–23].

Consistent with these findings, studies from Iringa and other Tanzanian regions have reported similar associations of LTFU with younger age and male gender, supporting the coexistence of individual-level risk factors and contextual service-delivery drivers [24]. Together, these observations suggest that disengagement from care reflects both patient-level characteristics and health-system organization, rather than a single underlying cause.

LTFU also varied by the year of the first recorded clinic visit, with higher proportions observed among individuals entering care in earlier calendar years. This pattern likely reflects cumulative exposure to disengagement over longer follow-up rather than differences in baseline risk. Lower attrition observed among more recent cohorts may be associated with programmatic improvements, including differentiated service delivery models, multi-month dispensing, and strengthened tracing systems [4,6,8,25,26]. Similar cohort patterns have been described in other sub-Saharan African settings, particularly among adolescents and young adults [4,19,24,27,28].

These findings suggest that pharmacy refill adherence could be incorporated into routine HIV monitoring as an objective and scalable indicator for early identification of patients at risk of disengagement. Combining adherence indicators with spatial analysis may enable programmes to prioritize districts and populations where disengagement is concentrated, supporting targeted retention interventions and sustained treatment effectiveness [1–3,5,6,25,29]. In conjunction with prior regional studies in Tanzania, the results support a differentiated and geographically responsive retention strategy informed by routinely collected adherence data.

## Limitations

This study relied on routinely collected programme data and is therefore subject to limitations inherent in secondary data sources. Because loss to follow-up (LTFU) was defined using visit-date gaps within a fixed observation window, the operational measure may capture a mixture of true disengagement from care, undocumented transfers between facilities, and delays in documentation. Missing or incomplete records may have affected the accuracy of some variables, and undocumented transfers between facilities may have resulted in misclassification of some individuals as lost to follow-up. Pharmacy refill adherence reflects medication pickup rather than actual medication ingestion and may therefore overestimate true adherence. Viral load measurements were unavailable for a subset of participants, limiting the evaluation of virological outcomes among those individuals. In addition, the database lacked detailed socioeconomic and behavioral variables, preventing assessment of certain patient-level determinants of disengagement. Despite these limitations, the large nationally representative cohort, consistent patterns observed across regions and subpopulations, and concordance with prior Tanzanian and international studies support the robustness and programmatic relevance of the findings.

## Conclusion

LFTU remains a substantial challenge for continuity of ART care in Tanzania and shows marked geographic variation across regions and districts. Pharmacy refill adherence was strongly associated with disengagement from care, with individuals demonstrating good adherence showing substantially lower odds of LTFU within routine programme data. Incorporating refill-based monitoring together with spatial analysis may enable earlier identification of individuals and areas at increased risk of attrition and support targeted retention interventions within HIV treatment programmes.

## Declarations

### Ethics approval and consent to participate

This study used routinely collected, de-identified programmatic data from the National Care and Treatment Clinic (CTC-2) database maintained by the National AIDS and Sexually Transmitted Infection Control Programme (NASHCoP) in Tanzania. Ethical approval for the secondary analysis of these data was obtained from the Muhimbili University of Health and Allied Sciences (MUHAS) Institutional Review Board. Permission to access and analyze the data was granted by NASHCoP. Individual informed consent was waived because the study involved a retrospective analysis of anonymized routine health data.

### Consent for publication

Not applicable. This manuscript does not contain identifiable individual-level data.

### Availability of data and materials

The data used in this study originate from the National AIDS and Sexually Transmitted Infections Control Programme (NASHCoP) and are not publicly available due to programmatic confidentiality restrictions. The data custodians may grant access upon reasonable request and with appropriate approvals.

## Authors’ contributions

MDL contributed to the study design, data preparation, and statistical analysis.

HM and LDV contributed to the data analysis and interpretation of the results.

RZS conceived the study, provided overall supervision, guided the analytical framework, critically interpreted the findings, and revised the manuscript accordingly.

RFM contributed to the clinical interpretation and contextualisation of the findings.

JM contributed to the methodological input and data interpretation.

VS and PN facilitated access to programmatic data, provided national programme oversight, and

contributed to the interpretation of results within the HIV programme context.

All the authors have reviewed and approved the final manuscript.

## Acknowledgements

The authors acknowledge the National AIDS and Sexually Transmitted Infections Control Programme (NASHCoP) for granting access to the CTC-2 database. We also thank the regional and district HIV programme teams and healthcare workers across Tanzania for their roles in routine data collection and programme implementation.

## Disclaimer

The views expressed in this article are those of the authors and do not necessarily reflect the official policies or positions of the National AIDS and Sexually Transmitted Infections Control Programme, Ministry of Health, Royal Society of Tropical Medicine and Hygiene or affiliated institutions.

## References

1. UNAIDS. Global HIV & AIDS statistics - Fact sheet. 2024 [cited 22 Jan 2026]. Available: https://www.unaids.org/en/resources/fact-sheet

2. Carter A, Zhang M, Tram KH, Walters MK, Jahagirdar D, Brewer ED, et al. Global, regional, and national burden of HIV/AIDS, 1990-2021, and forecasts to 2050, for 204 countries and territories: the Global Burden of Disease Study 2021. Lancet HIV. 2024;11: e807-e822. doi:10.1016/S2352-3018(24)00212-1

3. Parker E, Judge MA, Macete E, Nhampossa T, Dorward J, Langa DC, et al. HIV infection in Eastern and Southern Africa: Highest burden, largest challenges, greatest potential. South Afr J HIV Med. 2021;22. doi:10.4102/sajhivmed.v22i1.1237

4. Kilapilo MS, Sangeda RZ, Bwire GM, Sambayi GL, Mosha IH, Killewo J. Adherence to Antiretroviral Therapy and Associated Factors Among People Living With HIV Following the Introduction of Dolutegravir Based Regimens in Dar es Salaam, Tanzania. Journal of the International Association of Providers of AIDS Care (JIAPAC). 2022;21: 232595822210845. doi:10.1177/23259582221084543

5. United Republic of Tanzania. Tanzania HIV Impact Survey 2022-2023. 2024 [cited 18 Dec 2025]. Available: https://www.nbs.go.tz/nbs/takwimu/THIS2022-2023/THIS2022-2023_Summary_Sheet.pdf

6. UNAIDS. An ambitious treatment target to help end the AIDS epidemic. 2014 [cited 15 Nov 2020]. Available: https://www.unaids.org/en/resources/909090

7. Grossberg R, Gross R. Use of pharmacy refill data as a measure of antiretroviral adherence. Curr HIV/AIDS Rep. 2007;4: 187–91. Available: http://www.ncbi.nlm.nih.gov/pubmed/18366950

8. Panel on Antiretroviral Guidelines for Adults and Adolescents. Guidelines for the Use of Antiretroviral Agents in Adults and Adolescents With HIV. 2025 [cited 10 Jan 2026]. Available: https://clinicalinfo.hiv.gov/sites/default/files/guidelines/documents/adult-adolescent-arv/guidelines-adult-adolescent-arv.pdf#page=3.49

9. Sangeda RZ, Mosha F, Prosperi M, Aboud S, Vercauteren J, Camacho RJ, et al. Pharmacy refill adherence outperforms self-reported methods in predicting HIV therapy outcome in resource-limited settings. BMC Public Health. 2014;14: 1035. doi:10.1186/1471-2458-14-1035

10. Amour M, Sangeda RZ, Kidenya B, Balandya E, Mmbaga BT, Machumi L, et al. Adherence to Antiretroviral Therapy by Medication Possession Ratio and Virological Suppression among Adolescents and Young Adults Living with HIV in Dar es Salaam, Tanzania. Tropical Medicine and Infectious Disease 2022, Vol 7, Page 52. 2022;7: 52. doi:10.3390/TROPICALMED7040052

11. Martin D, Luz PM, Lake JE, Clark JL, Campos DP, Veloso VG, et al. Pharmacy refill data can be used to predict virologic failure for patients on antiretroviral therapy in Brazil. J Int AIDS Soc. 2017;20. doi:10.7448/IAS.20.1.21405

12. Messou E, Kouakou M, Gabillard D, Gouessé P, Koné M, Tchehy A, et al. Medication Possession Ratio: Predicting and Decreasing Loss to Follow-Up in Antiretroviral Treatment Programs in Côte d’Ivoire. JAIDS Journal of Acquired Immune Deficiency Syndromes. 2011;57: S34–S39. doi:10.1097/QAI.0b013e3182208003

13. Hong SY, Winston A, Mutenda N, Hamunime N, Roy T, Wanke C, et al. Predictors of loss to follow-up from HIV antiretroviral therapy in Namibia. Kufa T, editor. PLoS One. 2022;17: e0266438. doi:10.1371/journal.pone.0266438

14. Mushy SE, Mtisi E, Mboggo E, Mkawe S, Yahya-Malima KI, Ndega J, et al. Predictors of the observed high prevalence of loss to follow-up in ART-experienced adult PLHIV: a retrospective longitudinal cohort study in the Tanga Region, Tanzania. BMC Infect Dis. 2023;23: 92. doi:10.1186/s12879-023-08063-9

15. Siril HN, Kaaya SF, Smith Fawzi MK, Mtisi E, Somba M, Kilewo J, et al. CLINICAL outcomes and loss to follow-up among people living with HIV participating in the NAMWEZA intervention in Dar es Salaam, Tanzania: a prospective cohort study. AIDS Res Ther. 2017;14: 18. doi:10.1186/s12981-017-0145-z

16. Kalulo MB, Sangeda RZ, Mwakyomo J, Sangeda GR, Sambu V, Njau P. Utilizing pharmacy refill data to predict loss to follow-up among people living with HIV in Manyara region of Tanzania. 2026. doi:10.64898/2026.02.24.26347034

17. Mushi H, Lugoba MD, Sangeda RZ, Mutagonda RF, Mwakyomo J, Musiba G, et al. Predictors of loss to follow-up among patients receiving antiretroviral therapy in Njombe Region, Tanzania, 2017-2021. 2026. doi:10.64898/2026.02.28.26347333

18. Ochieng-Ooko V, Ochieng D, Sidle JE, Holdsworth M, Wools-Kaloustian K, Siika AM, et al. Influence of gender on loss to follow-up in a large HIV treatment programme in western Kenya. Bull World Health Organ. 2010;88: 681–688. doi:10.2471/BLT.09.064329

19. Kranzer K, Bradley J, Musaazi J, Nyathi M, Gunguwo H, Ndebele W, et al. Loss to follow up among children and adolescents growing up with HIV infection: age really matters. J Int AIDS Soc. 2017;20. doi:10.7448/IAS.20.1.21737

20. Minja AA, Larson E, Aloyce Z, Araya R, Kaale A, Kaaya SF, et al. Burden of HIV-related stigma and associated factors among women living with depression accessing PMTCT services in Dar es Salaam, Tanzania. AIDS Care. 2022;34: 1572–1579. doi:10.1080/09540121.2022.2050174

21. Kalinjuma A V, Glass TR, Weisser M, Myeya SJ, Kasuga B, Kisung’a Y, et al. Prospective assessment of loss to follow up: incidence and associated factors in a cohort of HIV positive adults in rural Tanzania. J Int AIDS Soc. 2020;23. doi:10.1002/jia2.25460

22. Manyanga VP, Mwakyomo J, Mushi H, Lugoba MD, Sambu V, Mutagonda RF, et al. Geospatial Quantification of Antiretroviral Therapy Consumption in Tanzania (2017-2021) Using the WHO Defined Daily Dose Methodology. 2025. doi:10.64898/2025.12.06.25341745

23. Machumi LC, Mtisi E, Andrew I, Sando D, Mkali H, Liu E, et al. Who are they? Identifying risk factors of loss to follow up among HIV+ patients on care and treatment in Dar es Salaam. BMC Infect Dis. 2014;14: P78. doi:10.1186/1471-2334-14-S2-P78

24. Barongo A, Kwesigabo G. Incidence and Determinants of Lost to Follow Up Among Adults Living with HIV/AIDS on Antiretroviral Therapy in Iringa Region, Tanzania. Int J Innov Sci Res Technol. 2025; 3869–3881. doi:10.38124/ijisrt/25apr1931

25. World Health Organization. Consolidated guidelines on the use of antiretroviral drugs for treating and preventing HIV infection: recommendations for a public health approach, 2nd ed. World Health Organization. 2016 [cited 15 Nov 2020]. Available: https://apps.who.int/iris/handle/10665/208825

26. Tesha E-D, Kishimba R, Njau P, Revocutus B, Mmbaga E. Predictors of loss to follow up from antiretroviral therapy among adolescents with HIV/AIDS in Tanzania. Zanoni BC, editor. PLoS One. 2022;17: e0268825. doi:10.1371/journal.pone.0268825

27. Bekolo CE, Webster J, Batenganya M, Sume GE, Kollo B. Trends in mortality and loss to follow-up in HIV care at the Nkongsamba Regional hospital, Cameroon. BMC Res Notes. 2013;6: 512. doi:10.1186/1756-0500-6-512

28. Hønge BL, Jespersen S, Nordentoft PB, Medina C, da Silva D, da Silva ZJ, et al. Loss to follow-up occurs at all stages in the diagnostic and follow-up period among HIV-infected patients in Guinea-Bissau: a 7-year retrospective cohort study. BMJ Open. 2013;3: e003499. doi:10.1136/bmjopen-2013-003499

29. Aliyu A, Adelekan B, Andrew N, Ekong E, Dapiap S, Murtala-Ibrahim F, et al. Predictors of loss to follow-up in art experienced patients in Nigeria: a 13 year review (2004-2017). AIDS Res Ther. 2019;16: 30. doi:10.1186/s12981-019-0241-3

