## Supplementary Materials for "Geographic variation in loss to follow-up from HIV care in Tanzania and its association with pharmacy refill adherence in routine programme data"

### Supplementary Material

### Supplementary Figures

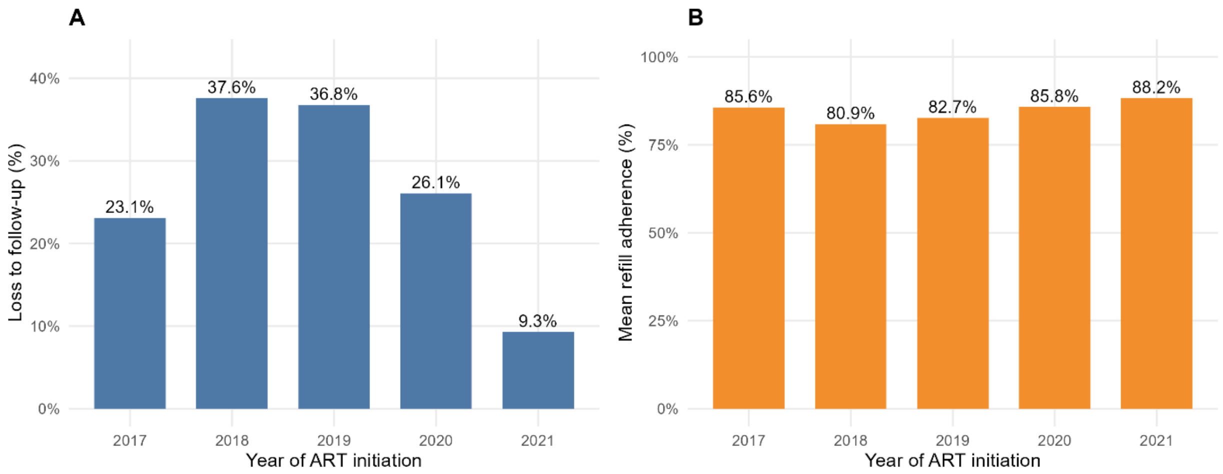

**Supplementary Figure S1: Loss to follow-up by year of ART initiation in Tanzania, 2017-2021**

**Panel A.** Proportion of people living with HIV classified as lost to follow-up (LTFU), defined as no recorded clinic visit for ≥180 days after the last scheduled appointment, stratified by calendar year of first recorded clinic visit in the registry extract.

**Panel B.** Number of individuals included in each cohort year of first recorded clinic visit. Earlier cohorts show higher proportions of LTFU, while more recent cohorts show lower proportions, reflecting shorter cumulative follow-up time.

Percentages were calculated separately for each cohort year. Analyses were restricted to mainland Tanzania.

### Supplementary Tables

**Supplementary Table S1: Regional distribution of individuals included in the analytic cohort derived from the Tanzania National HIV Care Registry (NASHCoP snapshot), 2017-2021 (N = 52,828).**

| Region | Number of PLHIV | Percent |
|---|---|---|
| Dar es Salaam | 6866 | 13 |
| Mbeya | 4010 | 7.6 |
| Mwanza | 4000 | 7.6 |
| Kagera | 3110 | 5.9 |
| Tabora | 2737 | 5.2 |
| Shinyanga | 2631 | 5 |
| Geita | 2625 | 5 |
| Morogoro | 2322 | 4.4 |
| Iringa | 2277 | 4.3 |
| Njombe | 2265 | 4.3 |
| Tanga | 1979 | 3.7 |
| Ruvuma | 1973 | 3.7 |
| Pwani | 1780 | 3.4 |
| Mara | 1758 | 3.3 |
| Songwe | 1437 | 2.7 |
| Dodoma | 1399 | 2.6 |
| Kilimanjaro | 1214 | 2.3 |
| Simiyu | 1207 | 2.3 |
| Mtwara | 1138 | 2.2 |
| Arusha | 1127 | 2.1 |
| Rukwa | 1052 | 2 |
| Katavi | 920 | 1.7 |
| Kigoma | 816 | 1.5 |
| Lindi | 811 | 1.5 |
| Singida | 780 | 1.5 |
| Manyara | 594 | 1.1 |

**Supplementary Table S2: District-level distribution of pharmacy refill adherence and loss to follow-up (LTFU) (≥180 days) among people living with HIV (PLHIV) receiving ART in mainland Tanzania, derived from the Tanzania National HIV Care Registry (NASHCoP snapshot), 2017-2021 (N = 52,828).**

| Region | District | Number of PLHIV | Mean refill adherence % | Poor refill adherence % | LTFU % |
|---|---|---|---|---|---|
| Arusha | Arumeru | 169 | 89.2 | 27.6 | 26.6 |
| Arusha | Arusha | 731 | 84.6 | 37.5 | 28.5 |
| Arusha | Karatu | 90 | 92.8 | 13.3 | 22.2 |
| Arusha | Longido | 41 | 77 | 47.5 | 39 |
| Arusha | Monduli | 71 | 77.9 | 49.3 | 40.8 |
| Arusha | Ngorongoro | 25 | 70.1 | 64 | 52 |
| Dar es Salaam | Ilala | 2372 | 87.4 | 27.4 | 36.6 |
| Dar es Salaam | Kigamboni | 297 | 86.8 | 25.4 | 37.4 |
| Dar es Salaam | Kinondoni | 1601 | 88.4 | 23.8 | 32.9 |
| Dar es Salaam | Temeke | 1819 | 86.3 | 29.7 | 26.9 |
| Dar es Salaam | Ubungo | 777 | 87.9 | 24.7 | 36.8 |
| Dodoma | Bahi | 122 | 78.6 | 43.3 | 40.2 |
| Dodoma | Chamwino | 154 | 88.7 | 25.5 | 20.1 |
| Dodoma | Chemba | 31 | 83.8 | 50 | 25.8 |
| Dodoma | Dodoma | 638 | 85.4 | 36.4 | 28.4 |
| Dodoma | Kondoa | 109 | 85.2 | 37.6 | 42.2 |
| Dodoma | Kongwa | 196 | 85.3 | 36.4 | 32.1 |
| Dodoma | Mpwapwa | 149 | 88.6 | 23 | 19.5 |
| Geita | Bukombe | 347 | 85.9 | 24.3 | 28.8 |
| Geita | Chato | 634 | 84.3 | 34 | 32.5 |
| Geita | Geita | 1191 | 84 | 34.3 | 35.5 |
| Geita | Mbogwe | 313 | 84.7 | 32.1 | 28.4 |
| Geita | Nyang'hwale | 140 | 87.1 | 25.7 | 28.6 |
| Iringa | Iringa | 1047 | 90 | 20.3 | 20.6 |
| Iringa | Kilolo | 252 | 87.3 | 29.5 | 29 |
| Iringa | Mafinga | 318 | 88.2 | 25.8 | 17.3 |
| Iringa | Mufindi | 660 | 89.6 | 19.4 | 18 |
| Kagera | Biharamulo | 263 | 83.4 | 34.5 | 30 |
| Kagera | Bukoba | 918 | 87.3 | 24.7 | 24.5 |
| Kagera | Karagwe | 316 | 83.6 | 36.5 | 24.4 |
| Kagera | Kyerwa | 323 | 81.9 | 43.4 | 25.4 |
| Kagera | Missenyi | 387 | 88 | 25.4 | 27.9 |
| Kagera | Muleba | 715 | 86.7 | 28.9 | 27.6 |
| Kagera | Ngara | 188 | 84.9 | 28.3 | 31.4 |

| | | | | | |
|---|---|---|---|---|---|
| Katavi | Mlele | 468 | 78.5 | 48.1 | 31.8 |
| Katavi | Mpanda | 452 | 83.9 | 35.1 | 33 |
| Kigoma | Buhigwe | 25 | 89.4 | 37.5 | 28 |
| Kigoma | Kakonko | 87 | 84.7 | 27.9 | 26.4 |
| Kigoma | Kasulu | 110 | 85.9 | 29.1 | 23.6 |
| Kigoma | Kibondo | 105 | 84.8 | 31.4 | 40 |
| Kigoma | Kigoma | 212 | 83.1 | 34 | 53.3 |
| Kigoma | Uvinza | 277 | 81.4 | 44 | 20.6 |
| Kilimanjaro | Hai | 121 | 81.4 | 42.1 | 27.3 |
| Kilimanjaro | Moshi | 662 | 87.6 | 27.9 | 23.3 |
| Kilimanjaro | Mwanga | 80 | 82.3 | 45 | 18.8 |
| Kilimanjaro | Rombo | 116 | 86.9 | 31 | 21.6 |
| Kilimanjaro | Same | 138 | 75.2 | 63 | 23.9 |
| Kilimanjaro | Siha | 97 | 83 | 39.6 | 30.9 |
| Lindi | Kilwa | 137 | 78.5 | 48.9 | 26.3 |
| Lindi | Lindi | 318 | 84.3 | 37.7 | 22.6 |
| Lindi | Liwale | 57 | 76.8 | 45.5 | 31.6 |
| Lindi | Nachingwea | 174 | 80.3 | 49.1 | 23.6 |
| Lindi | Ruangwa | 125 | 90.6 | 20.2 | 16.8 |
| Manyara | Babati | 211 | 86 | 31.4 | 26.1 |
| Manyara | Hanang | 50 | 87.5 | 32 | 32 |
| Manyara | Kiteto | 142 | 83.2 | 44.4 | 24.6 |
| Manyara | Mbulu | 69 | 86.2 | 33.3 | 18.8 |
| Manyara | Simanjiro | 122 | 76 | 55.7 | 48.4 |
| Mara | Bunda | 340 | 85.9 | 27.4 | 17.1 |
| Mara | Butiama | 140 | 83.4 | 40.3 | 28.6 |
| Mara | Musoma | 443 | 86.1 | 32.6 | 25.1 |
| Mara | Rorya | 518 | 81.8 | 48.6 | 16.6 |
| Mara | Serengeti | 128 | 86.2 | 36.5 | 20.3 |
| Mara | Tarime | 189 | 84.8 | 41.4 | 21.2 |
| Mbeya | Chunya | 419 | 79.8 | 55.9 | 24.6 |
| Mbeya | Kyela | 600 | 84.3 | 39.9 | 19 |
| Mbeya | Mbarali | 780 | 84.4 | 35.7 | 26 |
| Mbeya | Mbeya | 1637 | 84.7 | 36.2 | 27 |
| Mbeya | Rungwe | 574 | 83.2 | 43.7 | 19.3 |
| Morogoro | Gairo | 98 | 91 | 16.3 | 13.3 |
| Morogoro | Kilombero | 652 | 82.6 | 39.9 | 19.5 |
| Morogoro | Kilosa | 513 | 77.8 | 51.8 | 29 |
| Morogoro | Morogoro | 588 | 82.4 | 38.4 | 31.6 |
| Morogoro | Mvomero | 207 | 79.2 | 55.9 | 40.6 |
| Morogoro | Ulanga | 264 | 82.8 | 37.1 | 16.3 |
| Mtwara | Masasi | 435 | 85.5 | 33.7 | 23.2 |
| Mtwara | Mtwara | 333 | 79.7 | 47.4 | 25.8 |

| | | | | | |
|---|---|---|---|---|---|
| Mtwara | Nanyumbu | 88 | 74.4 | 58 | 25 |
| Mtwara | Newala | 148 | 82.3 | 46.2 | 21.6 |
| Mtwara | Tandahimba | 134 | 85 | 44.4 | 20.9 |
| Mwanza | Ilemela | 501 | 83.4 | 41.3 | 21 |
| Mwanza | Kwimba | 511 | 89.5 | 21.7 | 11.7 |
| Mwanza | Magu | 439 | 84.4 | 35.6 | 22.3 |
| Mwanza | Misungwi | 389 | 85.3 | 35.9 | 18.3 |
| Mwanza | Nyamagana | 995 | 89.8 | 20.6 | 21.8 |
| Mwanza | Sengerema | 802 | 86.6 | 30.3 | 18.7 |
| Mwanza | Ukerewe | 363 | 86.1 | 34.1 | 17.9 |
| Njombe | Ludewa | 323 | 87.7 | 26.6 | 29.1 |
| Njombe | Makete | 366 | 89.1 | 25 | 20.2 |
| Njombe | Njombe | 1218 | 85.1 | 33.8 | 32.9 |
| Njombe | Wanging'ombe | 358 | 86.2 | 31.2 | 49.2 |
| Pwani | Bagamoyo | 507 | 83.9 | 36.1 | 28.6 |
| Pwani | Kibaha | 453 | 83.7 | 32.7 | 31.3 |
| Pwani | Kisarawe | 169 | 88.3 | 25.7 | 29.6 |
| Pwani | Mafia | 46 | 71.6 | 57.8 | 37 |
| Pwani | Mkuranga | 302 | 85.2 | 34.8 | 22.8 |
| Pwani | Rufiji | 303 | 84.8 | 33.4 | 28.1 |
| Rukwa | Kalambo | 85 | 76.5 | 51.8 | 32.9 |
| Rukwa | Nkasi | 235 | 72.1 | 66.5 | 29.8 |
| Rukwa | Sumbawanga | 732 | 78.2 | 50.1 | 29 |
| Ruvuma | Mbinga | 484 | 84 | 43.3 | 21.1 |
| Ruvuma | Namtumbo | 185 | 78.5 | 51.1 | 27.6 |
| Ruvuma | Nyasa | 215 | 76.9 | 61.5 | 22.3 |
| Ruvuma | Songea | 836 | 85.7 | 35.4 | 23.1 |
| Ruvuma | Tunduru | 253 | 74.8 | 53.2 | 32.8 |
| Shinyanga | Kahama | 1540 | 80.9 | 40.2 | 28 |
| Shinyanga | Kishapu | 274 | 81.8 | 40.1 | 23.4 |
| Shinyanga | Shinyanga | 817 | 83.2 | 37.1 | 24.4 |
| Simiyu | Bariadi | 332 | 80.7 | 42.3 | 30.1 |
| Simiyu | Busega | 270 | 86.4 | 30.5 | 25.9 |
| Simiyu | Itilima | 144 | 84.2 | 38 | 20.1 |
| Simiyu | Maswa | 285 | 84.4 | 33.6 | 21.1 |
| Simiyu | Meatu | 176 | 88 | 30.1 | 28.4 |
| Singida | Ikungi | 102 | 77.1 | 59.8 | 19.6 |
| Singida | Iramba | 190 | 83.2 | 45 | 23.2 |
| Singida | Manyoni | 231 | 82.4 | 47.4 | 23.8 |
| Singida | Mkalama | 65 | 83.6 | 44.6 | 15.4 |
| Singida | Singida | 192 | 75.7 | 57.9 | 22.9 |
| Songwe | Ileje | 84 | 84.9 | 39.3 | 14.3 |
| Songwe | Mbozi | 640 | 86.4 | 32.4 | 18.8 |

| | | | | | |
|---|---|---|---|---|---|
| Songwe | Momba | 395 | 79.3 | 53.4 | 25.8 |
| Songwe | Songwe | 318 | 77.8 | 61.3 | 28.3 |
| Tabora | Igunga | 465 | 85.7 | 32.3 | 20.4 |
| Tabora | Kaliua | 434 | 83.7 | 41.1 | 27.4 |
| Tabora | Nzega | 701 | 85.5 | 31.1 | 25.5 |
| Tabora | Sikonge | 208 | 82.6 | 44 | 24.5 |
| Tabora | Tabora | 369 | 85.7 | 30.1 | 27.9 |
| Tabora | Urambo | 183 | 88 | 27.3 | 18 |
| Tabora | Uyui | 377 | 82.6 | 40.2 | 21.2 |
| Tanga | Handeni | 202 | 86.2 | 28.4 | 36.1 |
| Tanga | Kilindi | 94 | 81.3 | 46.8 | 36.2 |
| Tanga | Korogwe | 356 | 87.3 | 23.2 | 25.6 |
| Tanga | Lushoto | 209 | 88.6 | 23.7 | 25.8 |
| Tanga | Mkinga | 98 | 88.1 | 25.5 | 24.5 |
| Tanga | Muheza | 325 | 90.9 | 18.2 | 21.2 |
| Tanga | Pangani | 105 | 84.8 | 33.3 | 25.7 |
| Tanga | Tanga | 590 | 86.4 | 28.8 | 23.9 |

**Supplementary Table S3: District-level loss to follow-up (LTFU ≥180 days) by cohort year of first recorded clinic visit in the registry extract, mainland Tanzania, 2017-2021 (N = 52,828).**

District-level proportions of people living with HIV (PLHIV) classified as lost to follow-up (LTFU), defined as no recorded clinic visit for ≥180 days after the last scheduled appointment, stratified by calendar year of ART initiation (2017-2021).

| **Region** | **District** | **2017 %** | **2018 %** | **2019 %** | **2020 %** | **2021 %** |
|---|---|---|---|---|---|---|
| Arusha | Arumeru | 21.7 | 40 | 35.7 | 30.8 | 27.3 |
| Arusha | Arusha | 24 | 34.2 | 48.9 | 32.9 | 12.5 |
| Arusha | Karatu | 20.5 | 14.3 | 30.8 | 33.3 | 0 |
| Arusha | Longido | 21.1 | 62.5 | 42.9 | 66.7 | 0 |
| Arusha | Monduli | 25 | 64.3 | 40 | 58.3 | 33.3 |
| Arusha | Ngorongoro | 36.4 | 66.7 | 50 | 100 | |
| Dar es Salaam | Ilala | 31 | 55.4 | 54 | 37.1 | 12.1 |
| Dar es Salaam | Kigamboni | 25.8 | 58.7 | 66.1 | 25.9 | 5.9 |
| Dar es Salaam | Kinondoni | 29.4 | 45.7 | 48.6 | 25.9 | 12.9 |
| Dar es Salaam | Temeke | 22.4 | 38.6 | 39.7 | 31.5 | 13.3 |
| Dar es Salaam | Ubungo | 25.9 | 53.3 | 57.8 | 36.8 | 14.3 |
| Dodoma | Bahi | 45 | 55.6 | 26.7 | 35 | 11.1 |
| Dodoma | Chamwino | 25 | 35.7 | 25 | 13.9 | 0 |
| Dodoma | Chemba | 23.5 | 33.3 | 100 | 0 | 0 |

| | | | | | | |
|---|---|---|---|---|---|---|
| Dodoma | Dodoma | 22.3 | 28.4 | 44.4 | 41.6 | 16 |
| Dodoma | Kondoa | 38.3 | 41.2 | 50 | 66.7 | 30 |
| Dodoma | Kongwa | 22.5 | 40.7 | 54.2 | 43.9 | 6.7 |
| Dodoma | Mpwapwa | 15.6 | 36.4 | 26.1 | 17.2 | 0 |
| Geita | Bukombe | 24.6 | 31.9 | 41.2 | 34.1 | 5 |
| Geita | Chato | 27.4 | 44.3 | 40 | 33.3 | 16.9 |
| Geita | Geita | 24.1 | 44.2 | 45.4 | 45.9 | 18.3 |
| Geita | Mbogwe | 21.8 | 16.7 | 47.8 | 30.7 | 11.8 |
| Geita | Nyang'hwale | 22 | 35 | 42.4 | 30.8 | 0 |
| Iringa | Iringa | 16.9 | 31.2 | 28.9 | 22.3 | 4.7 |
| Iringa | Kilolo | 25.9 | 28.6 | 34 | 36.7 | 0 |
| Iringa | Mafinga | 15.4 | 20.6 | 29.2 | 5.9 | 6.2 |
| Iringa | Mufindi | 16.4 | 18.9 | 24.1 | 28.6 | 11.8 |
| Kagera | Biharamulo | 28 | 44.4 | 48.9 | 18.5 | 0 |
| Kagera | Bukoba | 22.5 | 34.5 | 34.7 | 14.5 | 4.1 |
| Kagera | Karagwe | 19.7 | 38.5 | 34.8 | 20.5 | 9.1 |
| Kagera | Kyerwa | 21.2 | 35.3 | 42.2 | 16.3 | 0 |
| Kagera | Missenyi | 18.9 | 41 | 52.1 | 7.5 | 0 |
| Kagera | Muleba | 24 | 41.7 | 35 | 23.5 | 0 |
| Kagera | Ngara | 16.7 | 40 | 62.2 | 38.5 | 10 |
| Katavi | Mlele | 27.4 | 44.9 | 40.7 | 26.3 | 0 |
| Katavi | Mpanda | 30 | 45.1 | 38.9 | 36.7 | 0 |
| Kigoma | Buhigwe | 25 | | 28.6 | 42.9 | 0 |
| Kigoma | Kakonko | 34.6 | 33.3 | 25 | 21.7 | 14.3 |
| Kigoma | Kasulu | 19.3 | 30.8 | 60 | 13.3 | 0 |
| Kigoma | Kibondo | 36.7 | 57.1 | 57.1 | 33.3 | 0 |
| Kigoma | Kigoma | 54.7 | 53.8 | 51.4 | 51.7 | 40 |
| Kigoma | Uvinza | 16.4 | 44 | 25 | 16 | 7.7 |
| Kilimanjaro | Hai | 19.1 | 56.2 | 31.2 | 33.3 | 16.7 |
| Kilimanjaro | Moshi | 18.4 | 41.2 | 38 | 33.3 | 3.8 |
| Kilimanjaro | Mwanga | 20 | 25 | 23.1 | 10 | 0 |
| Kilimanjaro | Rombo | 17.8 | 41.7 | 30 | 20 | 18.2 |
| Kilimanjaro | Same | 21.9 | 36 | 36.8 | 13 | 0 |
| Kilimanjaro | Siha | 20 | 80 | 40 | 40 | 50 |
| Lindi | Kilwa | 23.7 | 40 | 26.7 | 26.3 | 14.3 |
| Lindi | Lindi | 13.8 | 37.5 | 48.6 | 28.1 | 8.3 |
| Lindi | Liwale | 20.8 | 53.8 | 33.3 | 30 | 0 |
| Lindi | Nachingwea | 17.8 | 55.6 | 34.8 | 0 | 0 |
| Lindi | Ruangwa | 16.2 | 23.5 | 13.6 | 16.7 | 16.7 |
| Manyara | Babati | 26.5 | 29.6 | 40 | 19.4 | 0 |
| Manyara | Hanang | 22.6 | 25 | 54.5 | 50 | |
| Manyara | Kiteto | 26.4 | 20 | 25 | 22.2 | 20 |
| Manyara | Mbulu | 17.2 | 11.1 | 23.5 | 33.3 | 0 |

| | | | | | | |
|---|---|---|---|---|---|---|
| Manyara | Simanjiro | 43.9 | 55.6 | 42.9 | 63.6 | 33.3 |
| Mara | Bunda | 13.3 | 21.4 | 34 | 13.8 | 4.8 |
| Mara | Butiama | 24.6 | 47.6 | 39.1 | 11.1 | 22.2 |
| Mara | Musoma | 19.8 | 34.6 | 32.1 | 31.9 | 10.3 |
| Mara | Rorya | 17.3 | 29.6 | 16.7 | 9.8 | 6.1 |
| Mara | Serengeti | 20.3 | 25 | 18.2 | 27.8 | 0 |
| Mara | Tarime | 17.1 | 54.5 | 27 | 2.9 | 23.1 |
| Mbeya | Chunya | 22 | 44.2 | 34.1 | 20 | 9.1 |
| Mbeya | Kyela | 16.5 | 24.7 | 28.1 | 23.3 | 4.5 |
| Mbeya | Mbarali | 20.9 | 39.4 | 34.9 | 35.9 | 18.5 |
| Mbeya | Mbeya | 24.7 | 36.3 | 35.9 | 27.1 | 13.8 |
| Mbeya | Rungwe | 17.9 | 27.3 | 27.1 | 20.8 | 0 |
| Morogoro | Gairo | 11.1 | 22.2 | 24 | 3.2 | 16.7 |
| Morogoro | Kilombero | 15.7 | 30.3 | 29.5 | 12.9 | 9.3 |
| Morogoro | Kilosa | 21.8 | 34.9 | 41.2 | 27.5 | 9.1 |
| Morogoro | Morogoro | 26.4 | 42.2 | 43.7 | 33.3 | 3.2 |
| Morogoro | Mvomero | 32.6 | 43.5 | 53.2 | 48.6 | 20 |
| Morogoro | Ulanga | 17 | 16 | 22 | 15.2 | 0 |
| Mtwara | Masasi | 20.3 | 31 | 27.8 | 23.7 | 28.6 |
| Mtwara | Mtwara | 21.2 | 35.8 | 33.3 | 29.7 | 11.1 |
| Mtwara | Nanyumbu | 29.6 | 28 | 20 | 28.6 | 0 |
| Mtwara | Newala | 15.2 | 40 | 43.8 | 37.5 | 0 |
| Mtwara | Tandahimba | 19.2 | 26.3 | 33.3 | 21.1 | 0 |
| Mwanza | Ilemela | 25.3 | 26.5 | 23.6 | 10.6 | 11.6 |
| Mwanza | Kwimba | 16 | 15.9 | 10.9 | 8.8 | 2 |
| Mwanza | Magu | 19.3 | 33.3 | 32.8 | 16.7 | 0 |
| Mwanza | Misungwi | 12.5 | 27.1 | 29.2 | 15.7 | 8.7 |
| Mwanza | Nyamagana | 21.1 | 31.6 | 24.8 | 17.6 | 11.3 |
| Mwanza | Sengerema | 20.1 | 22 | 21.5 | 15.4 | 2 |
| Mwanza | Ukerewe | 18.3 | 38.1 | 29 | 9.7 | 2.4 |
| Njombe | Ludewa | 27.6 | 19.4 | 43.8 | 46.4 | 7.7 |
| Njombe | Makete | 17.5 | 28.6 | 32.3 | 30 | 20 |
| Njombe | Njombe | 26.6 | 37.8 | 50.8 | 42.9 | 17.2 |
| Njombe | Wanging'ombe | 42.5 | 61.5 | 58.5 | 67.6 | 22.2 |
| Pwani | Bagamoyo | 23.8 | 44.2 | 38.5 | 23.7 | 18.5 |
| Pwani | Kibaha | 23.9 | 37.3 | 54.5 | 40.6 | 8 |
| Pwani | Kisarawe | 25 | 44.4 | 28.6 | 43.5 | 14.3 |
| Pwani | Mafia | 19.2 | 66.7 | 71.4 | 62.5 | 0 |
| Pwani | Mkuranga | 19.5 | 36.8 | 31.7 | 15 | 21.4 |
| Pwani | Rufiji | 21.5 | 34.4 | 46.6 | 32.4 | 6.2 |
| Rukwa | Kalambo | 27.5 | 50 | 50 | 30 | 33.3 |
| Rukwa | Nkasi | 24.1 | 54.5 | 33.3 | 28 | 18.2 |
| Rukwa | Sumbawanga | 27.3 | 40.2 | 32.1 | 29.4 | 14.3 |

| | | | | | | |
|---|---|---|---|---|---|---|
| Ruvuma | Mbinga | 17.4 | 29.4 | 40.4 | 21.6 | 5.4 |
| Ruvuma | Namtumbo | 30.9 | 43.8 | 18.8 | 26.1 | 0 |
| Ruvuma | Nyasa | 18.8 | 28.2 | 17.4 | 33.3 | 0 |
| Ruvuma | Songea | 22.1 | 29.2 | 37.3 | 15.5 | 12.8 |
| Ruvuma | Tunduru | 28.8 | 40.5 | 45 | 36 | 23.8 |
| Shinyanga | Kahama | 27.4 | 38.8 | 32 | 22.1 | 5.4 |
| Shinyanga | Kishapu | 23.8 | 28.6 | 33.3 | 14.7 | 12.5 |
| Shinyanga | Shinyanga | 20.6 | 37.3 | 37.8 | 21.5 | 9.1 |
| Simiyu | Bariadi | 24.4 | 40 | 39.6 | 40.4 | 5.3 |
| Simiyu | Busega | 27 | 38.9 | 26.8 | 21.2 | 0 |
| Simiyu | Itilima | 17.9 | 25 | 29.6 | 20 | 0 |
| Simiyu | Maswa | 18.8 | 26.3 | 28.3 | 33.3 | 3.7 |
| Simiyu | Meatu | 29.9 | 36 | 29.4 | 25 | 11.8 |
| Singida | Ikungi | 14.6 | 29.2 | 17.6 | 26.7 | 0 |
| Singida | Iramba | 20 | 41.4 | 33.3 | 23.8 | 5 |
| Singida | Manyoni | 23.9 | 24.3 | 27.3 | 27.5 | 0 |
| Singida | Mkalama | 6.1 | 33.3 | 42.9 | 25 | 0 |
| Singida | Singida | 13.1 | 41.7 | 47.6 | 25 | 18.8 |
| Songwe | Ileje | 20.4 | 10 | 0 | 0 | 0 |
| Songwe | Mbozi | 17.4 | 26.2 | 19.4 | 23.2 | 13.3 |
| Songwe | Momba | 29.6 | 26.8 | 25.8 | 25.5 | 5.6 |
| Songwe | Songwe | 28.8 | 47.2 | 28.6 | 9.1 | 18.8 |
| Tabora | Igunga | 20.9 | 31.2 | 19.7 | 18.3 | 3.8 |
| Tabora | Kaliua | 29.2 | 40.4 | 36.5 | 18.5 | 6.5 |
| Tabora | Nzega | 21.5 | 41.7 | 24.1 | 31.5 | 7.9 |
| Tabora | Sikonge | 20.6 | 23.8 | 36.6 | 27.5 | 0 |
| Tabora | Tabora | 21.8 | 41.9 | 44.4 | 31.4 | 15.8 |
| Tabora | Urambo | 23 | 20 | 17.9 | 9.1 | 6.7 |
| Tabora | Uyui | 19.7 | 37.9 | 25 | 16.7 | 4.9 |
| Tanga | Handeni | 27.3 | 50 | 53.1 | 37.8 | 12.5 |
| Tanga | Kilindi | 24.2 | 40 | 50 | 45 | 20 |
| Tanga | Korogwe | 21.4 | 40.9 | 27.8 | 34.4 | 7.1 |
| Tanga | Lushoto | 13.6 | 44.4 | 35.3 | 41.9 | 0 |
| Tanga | Mkinga | 20 | 31.2 | 45.5 | 27.8 | 0 |
| Tanga | Muheza | 22.4 | 25 | 50 | 14.3 | 9.5 |
| Tanga | Pangani | 22.9 | 50 | 46.2 | 17.9 | 0 |
| Tanga | Tanga | 18.5 | 50 | 27.9 | 32.4 | 16.7 |

For each region, the first row shows the regional aggregate across all districts, and the subsequent rows show the individual districts within that region. Blank cells indicate that no eligible observations were available for that district-year cohort. The analyses were restricted to mainland Tanzania. Values are district-specific LTFU percentages within each ART initiation year (LTFU count divided by the number of PLHIV in the initiation cohort of that year). District aggregate values across years are not shown because the overall district LTFU is weighted by cohort size and reported elsewhere.

**Supplementary Table S4: Adjusted association between region of care and loss to follow-up (LTFU ≥180 days) among PLHIV receiving ART in mainland Tanzania, 2017-2021 (N = 52,828).**

Outcome**:** Loss to follow-up (LTFU), defined as ≥180 days without a recorded clinic visit after the last scheduled appointment.

| Region | aOR | 95% CI | p-value |
|---|---|---|---|
| Arusha | 1.70 | 1.45-1.99 | <0.001 |
| Dar es Salaam | 2.29 | 2.08-2.53 | <0.001 |
| Dodoma | 1.75 | 1.51-2.02 | <0.001 |
| Geita | 2.11 | 1.87-2.37 | <0.001 |
| Iringa | 1.21 | 1.06-1.39 | 0.005 |
| Kagera | 1.52 | 1.35-1.71 | <0.001 |
| Katavi | 1.86 | 1.58-2.20 | <0.001 |
| Kigoma | 2.05 | 1.72-2.44 | <0.001 |
| Kilimanjaro | 1.31 | 1.11-1.53 | 0.001 |
| Lindi | 1.15 | 0.95-1.39 | 0.139 |
| Manyara | 1.77 | 1.45-2.16 | <0.001 |
| Mara | 1.04 | 0.90-1.20 | 0.625 |
| Mbeya | 1.26 | 1.12-1.40 | <0.001 |
| Morogoro | 1.35 | 1.19-1.53 | <0.001 |
| Mtwara | 1.14 | 0.96-1.35 | 0.124 |
| Njombe | 2.20 | 1.95-2.49 | <0.001 |
| Pwani | 1.69 | 1.47-1.93 | <0.001 |
| Rukwa | 1.45 | 1.24-1.71 | <0.001 |
| Ruvuma | 1.15 | 1.00-1.31 | 0.050 |
| Shinyanga | 1.38 | 1.22-1.56 | <0.001 |
| Simiyu | 1.46 | 1.25-1.71 | <0.001 |
| Singida | 0.99 | 0.82-1.20 | 0.927 |

| | | | |
|---|---|---|---|
| Songwe | 1.05 | 0.90-1.22 | 0.525 |
| Tabora | 1.28 | 1.13-1.45 | <0.001 |
| Tanga | 1.65 | 1.45-1.89 | <0.001 |

Adjusted odds ratios (aORs) were estimated using a multivariable logistic regression model including pharmacy refill adherence category, age, gender, marital status, age group, and region. Arusha was used as the reference region as it was the omitted category in model parameterization.
